# Accessibility and regional disparities in nationwide 24-hour home medical care: A quantitative evaluation using the enhanced two-step floating catchment area method

**DOI:** 10.64898/2026.04.18.26351162

**Authors:** Yuki Egashira, Ryo Watanabe

**Author notes:** Correspondence author (YE). Y.E. designed the study, the main conceptual ideas, and the proof outline. Y.E. collected the data. Y.E. interpreted the results and worked on the manuscript. R.W. supervised the project. All authors discussed the results and commented on the manuscript.

## Abstract

With Japan’s rapidly aging population, demand for home healthcare is projected to increase by 62% by 2040. This study quantitatively evaluated accessibility to 24-hour home healthcare and regional disparities across all 335 secondary medical areas (SMAs) in Japan using the Enhanced Two-Step Floating Catchment Area (E2SFCA) method. We conducted a nationwide cross-sectional study analyzing approximately 430,000 population points at 500-meter mesh resolution. The E2SFCA integrated demand (age-adjusted population), supply (24-hour home care support clinics and hospitals), and transportation (road networks). Accessibility scores (ASs) and Gini coefficients were calculated for each SMA. Ward’s hierarchical cluster analysis classified regional types, and multiple regression based on the Penchansky and Thomas five-dimensional access framework identified factors associated with the median AS (ASM) and Gini coefficient. The median ASM was 45.71 (0.00–153.49), and the median Gini coefficient was 0.33 (0.06–0.93). Cluster analysis identified six types ranked by descending ASM, from C1 (high access, equitable; n = 48) to C6 (access desert; n = 23). C6 had a median ASM of 0.00 and Gini coefficient of 0.74, indicating virtually no access within a 30-minute catchment. Home-visit standardized claim ratios, used as external validation, declined monotonically from C1 (125.6) to C6 (17.6). For ASM, 24-hour visiting nursing stations (β = +0.369) and clinic physicians (β = +0.342) showed the strongest positive associations, with non-residential area negatively associated (β = −0.273). For the Gini coefficient, non-residential area showed the strongest positive association (β = +0.523). Taxable income per taxpayer was not significantly associated with either outcome. Non-residential area was associated with both lower accessibility and greater intra-regional inequality, suggesting that geographic constraints may limit the effectiveness of resource investment alone. Uniform nationwide implementation of policies shifting care from long-term care beds to home healthcare may not be feasible; region-specific approaches considering geographic characteristics are necessary.

## Introduction

The World Health Organization (WHO) global strategy on people-centered and integrated health services, published in 2015, advocates a shift from traditional hospital-centered healthcare systems to patient-centered models worldwide in response to increasing healthcare costs associated with longevity and chronic diseases [1]. Within this framework, home healthcare addresses transitional care needs despite challenges related to coordination and reliance on informal caregivers, making policy enhancement an urgent priority. New challenges have emerged as a result of population aging, which is transforming healthcare systems. WHO projections indicate that the global population aged 60 years and older will double by 2050, suggesting that this demographic shift may introduce novel challenges to healthcare resource allocation [2]. Rapid population aging is a common trend in Asian countries, including Japan, which is one of the world’s most aged societies. According to the “Report of the Review Committee on the New Regional Healthcare Vision,” published in December 2024, the demand for home healthcare is projected to increase by 62% from 2022 to 2040, making the development of 24-hour home healthcare delivery systems increasingly important [3].

Penchansky and Thomas proposed a five-dimensional model of healthcare access—Availability, Accessibility, Affordability, Accommodation, and Acceptability—which has been widely applied in health services research [4]. Among these dimensions, geographic access (Accessibility) is a particularly important determinant for services such as home healthcare, for which direct provision at the patient’s residence is required. Systematic reviews of spatial accessibility research indicate that although studies on primary care are abundant, research specifically focusing on home healthcare remains limited.

The Enhanced Two-Step Floating Catchment Area (E2SFCA) method is widely used for the quantitative evaluation of healthcare accessibility [5]. This method has the advantage of integrating demand, supply, and transportation networks and has been applied in studies of primary care and elderly care in countries such as China, Australia, and Canada [6–8]. Access barriers in rural areas and among older populations—including long travel distances and poor road conditions—have been reported internationally as factors that inhibit home healthcare utilization, underscoring the need for methods to visualize and quantify geographic disparities in accessibility [9].

In Japan, healthcare planning typically relies on population-based facility ratios and supply–demand analyses at the secondary medical area (SMA) level. However, geographic access indicators reflecting the travel required for physicians to visit patients’ homes are not incorporated into healthcare planning evaluation frameworks [10]. Although domestic studies have used the 2SFCA method to analyze visiting nursing accessibility in Hokkaido and hospital access in Tochigi Prefecture, no study has comprehensively evaluated 24-hour home healthcare across all 335 SMAs nationwide [11,12]. Furthermore, no study has quantified intra-regional inequality using Gini coefficients or validated its correspondence with actual healthcare utilization using standardized claim ratios (SCRs).

This study focused on 24-hour home healthcare and incorporated an analysis of regulatory factors based on the Penchansky and Thomas five-dimensional framework to provide evidence for policy development. The objectives were threefold: (1) to evaluate accessibility to 24-hour home healthcare across all 335 SMAs at a 500-meter mesh resolution using the E2SFCA method with age-adjusted demand and actual road network travel times; (2) to quantify intra-regional geographic disparities using Gini coefficients; and (3) to identify structural determinants of accessibility and intra-regional disparities through multiple regression analysis based on the Penchansky framework.

As a secondary objective, Ward’s hierarchical cluster analysis was performed to classify regional types, and external consistency with SCRs was examined.

## Materials and methods

### Study design and setting

This study evaluated all 335 SMAs in Japan using the E2SFCA method, integrating demand, supply, and transportation. The target facilities were 24-hour home care support clinics and hospitals (hereafter referred to as home care support clinics/hospitals). Prior research has demonstrated that the intensity of home healthcare provision contributes to the achievement of home-based end-of-life care [13,14].

### Data sources

#### Demand

Population data by sex and five-year age groups at a 500-meter mesh resolution, projected for 2025 based on the 2020 National Census, were used [15]. Age- and sex-specific claim counts for “home-visit medical care fees” from National Database (NDB) open data were used to adjust demand. The overall claim proportion was set to 1, and age- and sex-specific claim proportions were applied to the corresponding population at each 500-meter mesh point [16]. Meshes with a population of zero were excluded from the analysis.

#### Supply

Facilities registered as home care support clinics and hospitals were extracted from notification records submitted to the Regional Bureaus of Health and Welfare [17–24]. In Japan, facility categories are determined according to the number of medical personnel, such as full-time physicians. There are three categories of 24-hour home medical care facilities. In this study, supply capacity was assigned based on these categories as follows: (1) enhanced-function type (standalone): 3; (2) collaborative type: 1.5; and (3) conventional type: 1.0. These values reflect the minimum full-time physician requirements stipulated in the facility designation standards (e.g., the enhanced-function standalone type requires a minimum of three full-time physicians). These category-based weights were used as the supply capacity (Sj) in the E2SFCA calculation. The locations of medical institutions were determined by extracting addresses from notifications filed with the national government and converting them to geographic coordinates [25].

#### Transportation

Road network data for all of Japan were obtained from OpenStreetMap (OSM) [26]. OSM is an open-source map that can be freely edited by users and, although it is not an official government dataset, its use has been reported in multiple healthcare accessibility studies, supporting its reliability [8,27]. In this study, the road network was constructed as a network dataset, and travel times were calculated along actual road routes rather than straight-line distances.

Travel speeds were assigned according to road type and population concentration district (DID) status based on the National Road and Street Traffic Survey [28]. Travel was assumed to occur by automobile or on foot. DID geographic data were obtained from the National Land Information Download Site and were used to distinguish DID from non-DID areas [29]. The analysis was conducted under the assumption that vehicles were used exclusively on general roads and not on expressways.

### E2SFCA method

The E2SFCA method is a two-step approach that evaluates accessibility from both the supply and demand perspectives. In this study, the catchment area was divided into three travel-time zones at 10-minute intervals, and distance-decay weights were applied (0–10 min: 1.00 [W_1_], 10–20 min: 0.68 [W_2_], 20–30 min: 0.22 [W_3_]) [5].

#### Step 1: Supply-side evaluation

The demand population reachable within the catchment area from each facility j was aggregated using distance-based weights to calculate the supply-to-demand ratio (R_j_). This ratio represents the supply capacity of facility j relative to the surrounding demand.

R_j_ = S_j_ / (ΣP_k_×W_1_ + ΣP_k_×W_2_ + ΣP_k_×W_3_)
R_j_: Supply-to-demand ratio for facility j
S_j_: Supply capacity of facility j (category-based weight: 3, 1.5, or 1.0 in this study)
P_k_: Demand population at point k reachable from facility j (after age-specific adjustment)

#### Step 2: Demand-side evaluation

The supply-to-demand ratios (R_j_) of all facilities reachable from each point i were aggregated using distance-based weights to calculate AS_i_. This score represents the total accessible healthcare resources for residents at point i.

AS_i_ = (ΣR_j_×W_1_ + ΣR_j_×W_2_ + ΣR_j_×W_3_) × 1,000,000
AS_i_: Accessibility score at point i
R_j_: Supply-to-demand ratio of facility j reachable from point i (calculated in Step 1)
W_1_, W_2_, W_3_: Distance-decay weights
*Expressed per 1,000,000 population

#### Travel time calculation

Travel times were calculated using the Network Analyst extension in ArcGIS Pro 3.30. Road network data obtained from OSM were constructed as a network dataset, with travel speeds assigned according to road type.

### Quantification of regional disparities

Gini coefficients were calculated to quantify intra-regional disparities within each SMA. The Gini coefficient is widely used in economics as a measure of income distribution inequality and has also been applied as an indicator of disparities in healthcare access research using the E2SFCA method [6,30]. Values range from 0 to 1, with values closer to 0 indicating smaller disparities and values closer to 1 indicating greater disparities.

### Cluster analysis

Median AS (ASM) and Gini coefficients were standardized, and Ward’s hierarchical cluster analysis using Euclidean distance was performed. The optimal number of clusters was determined using the Calinski–Harabasz (CH) index (comparing k = 2–6) [31]. Descriptive comparisons across clusters were conducted using medians and interquartile ranges, and external consistency with home-visit SCRs was examined. Each cluster was labeled according to the combination of ASM and Gini coefficient levels.

### Statistical analysis

Descriptive statistics were calculated, and multiple regression analyses were performed with ASM and Gini coefficients as dependent variables. Variables were z-standardized, and standardized coefficients were estimated. Explanatory variables were pre-specified based on the Penchansky and Thomas five-dimensional access framework (Availability, Accessibility, Affordability, and Demand; Accommodation and Acceptability were excluded because of the difficulty of quantification using area-level aggregate data) [4]. Multicollinearity was assessed using variance inflation factors (VIFs < 10 for all variables). All medical resource variables were expressed per 100,000 population, proportions as percentages, and income in thousands of yen.

As a subgroup analysis, SCRs for home visits (FY 2022) were analyzed by cluster. SCR is an age-adjusted claim ratio standardized to the national average (= 100) [32]. Geographic analyses were conducted using ArcGIS Pro 3.30, and statistical analyses were performed using Stata 18.0 SE.

### Ethical considerations

This study was a secondary analysis that used only publicly available data and was therefore exempt from ethical review.

## Results

### Descriptive statistics

The accessibility scores (AS) ranged from 0.00 to 7,057.68. Gini coefficients ranged from 0.06 to 0.93. The total number of mesh points analyzed was 428,100. Table 1 presents the results for accessibility and regional disparities in 24-hour home healthcare across all 335 SMAs. The ASM was 45.71, and the median Gini coefficient was 0.33.

**Table 1.**
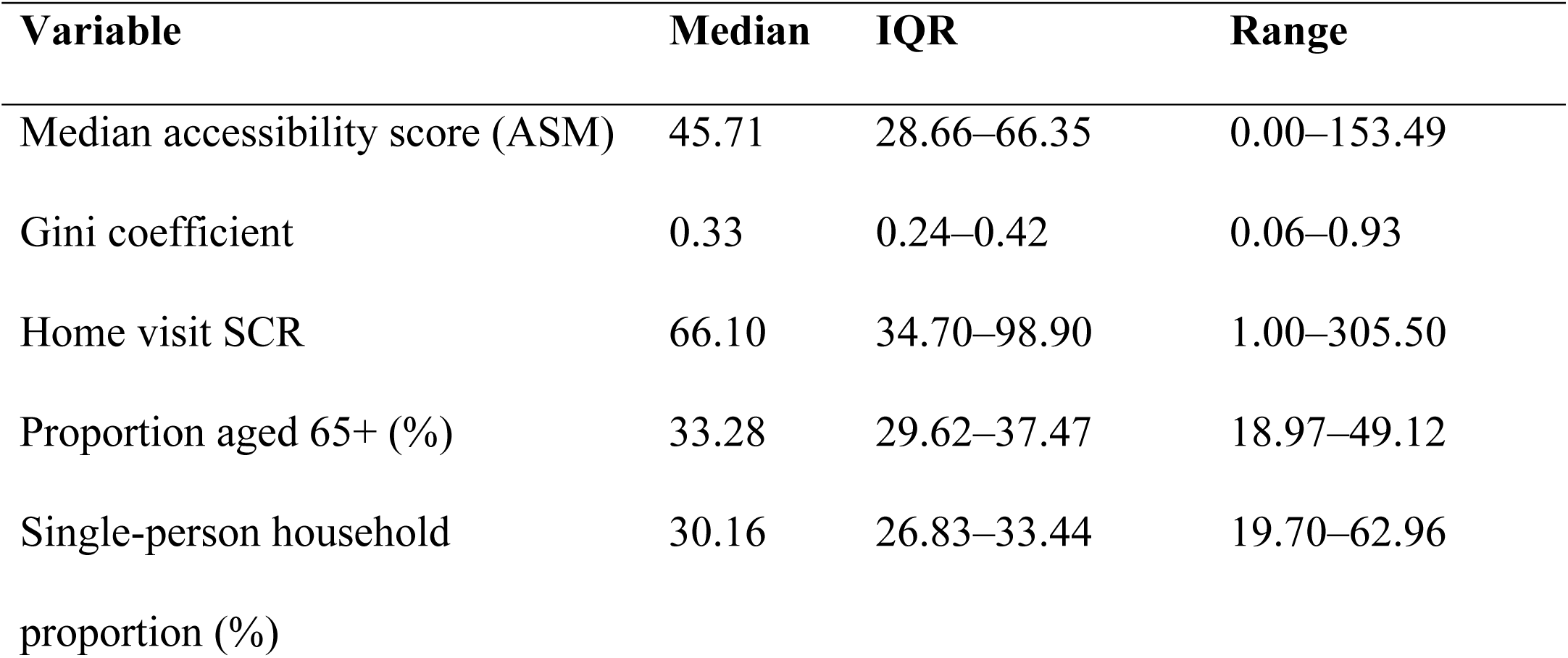
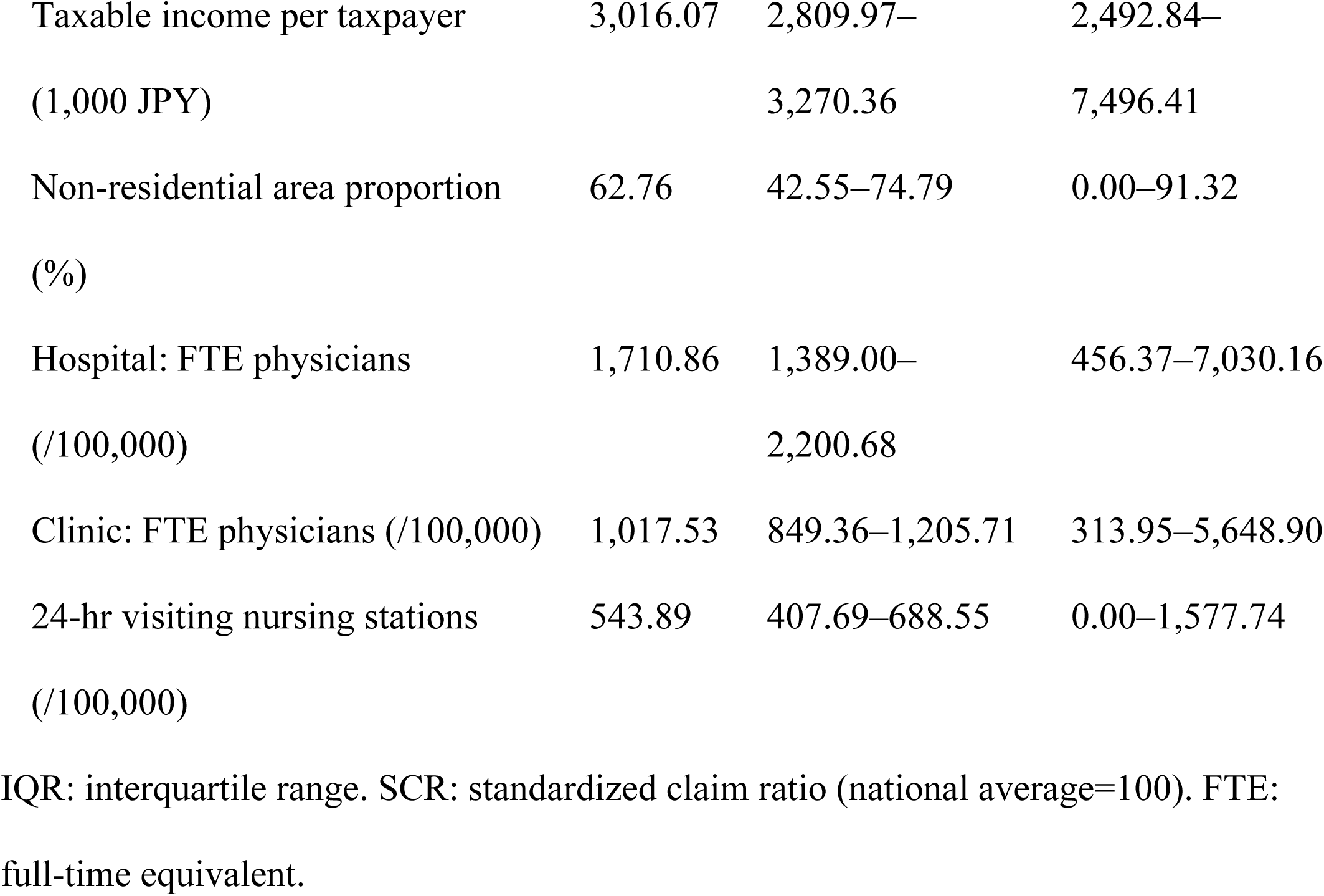
Descriptive statistics (n=335 secondary medical areas).

| Variable | Median | IQR | Range |
| --- | --- | --- | --- |
| Median accessibility score (ASM) | 45.71 | 28.66–66.35 | 0.00–153.49 |
| Gini coefficient | 0.33 | 0.24–0.42 | 0.06–0.93 |
| Home visit SCR | 66.10 | 34.70–98.90 | 1.00–305.50 |
| Proportion aged 65+ (%) | 33.28 | 29.62–37.47 | 18.97–49.12 |
| Single-person household proportion (%) | 30.16 | 26.83–33.44 | 19.70–62.96 |
| Taxable income per taxpayer<br>(1,000 JPY) | 3,016.07 | 2,809.97–<br>3,270.36 | 2,492.84–<br>7,496.41 |
| Non-residential area proportion<br>(%) | 62.76 | 42.55–74.79 | 0.00–91.32 |
| Hospital: FTE physicians<br>(/100,000) | 1,710.86 | 1,389.00–<br>2,200.68 | 456.37–7,030.16 |
| Clinic: FTE physicians (/100,000) | 1,017.53 | 849.36–1,205.71 | 313.95–5,648.90 |
| 24-hr visiting nursing stations<br>(/100,000) | 543.89 | 407.69–688.55 | 0.00–1,577.74 |

IQR: interquartile range. SCR: standardized claim ratio (national average=100). FTE: full-time equivalent.

The geographic distribution of ASM is shown in Fig 1. ASM was higher in metropolitan areas and lower in the Hokkaido and Tohoku regions.

**Fig 1.**
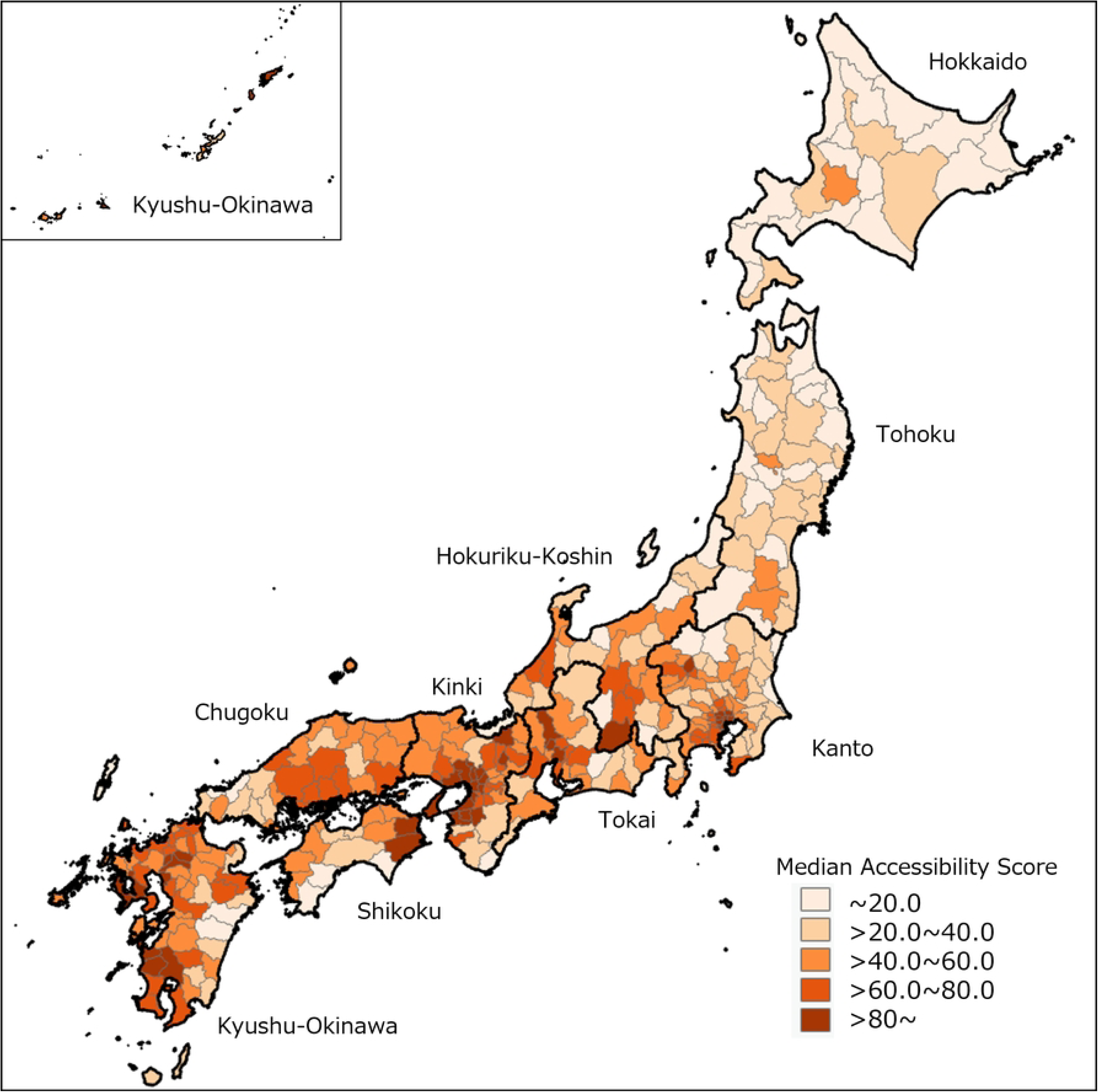
Geographic distribution of median accessibility scores (ASM) across 335 secondary medical areas.

Higher values indicate greater accessibility to 24-hour home healthcare.

The geographic distribution of Gini coefficients is shown in Fig 2. Gini coefficients were higher in the Hokkaido and Tohoku regions and lower in metropolitan areas.

**Fig 2.**
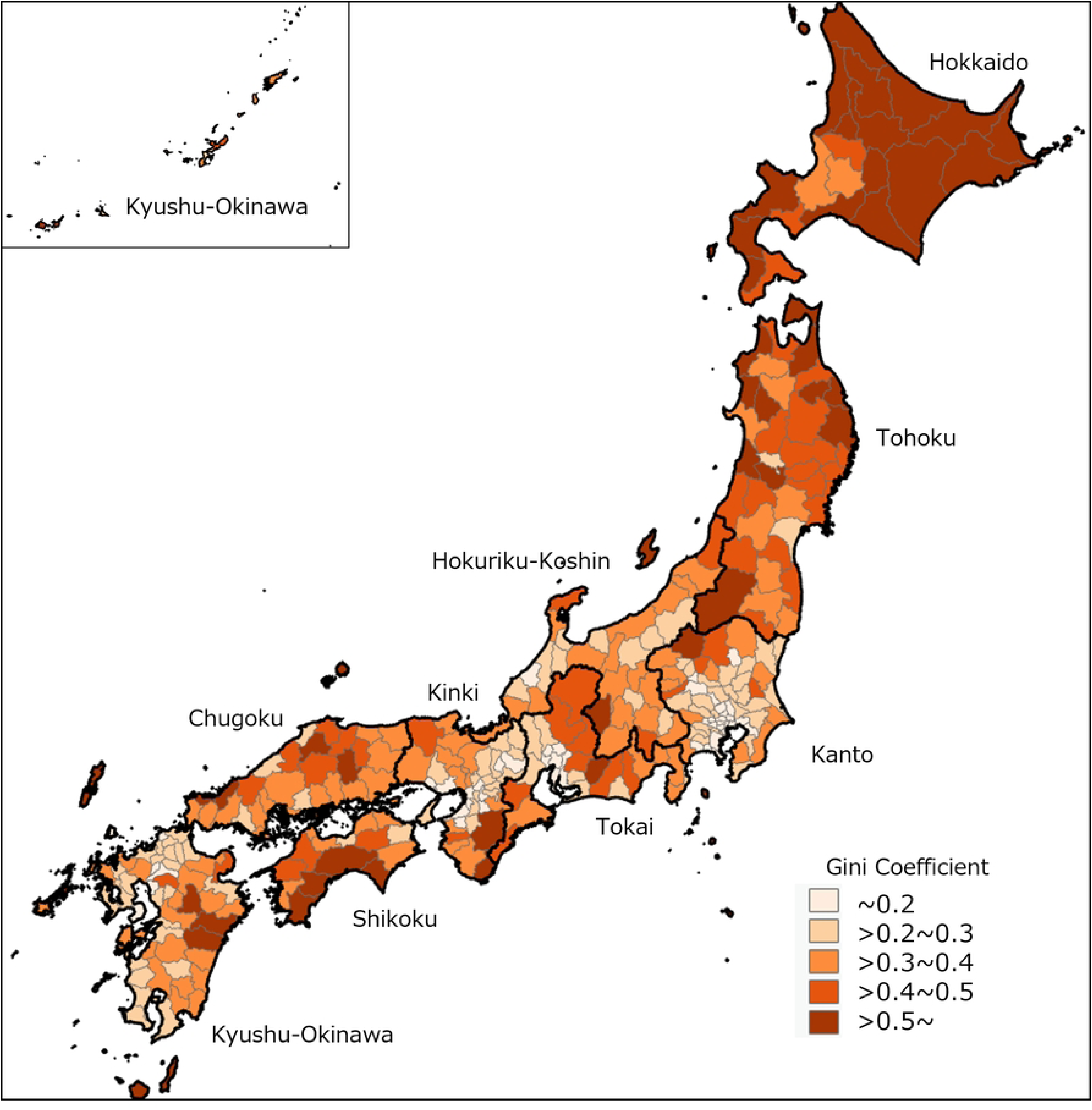
Geographic distribution of Gini coefficients across 335 secondary medical areas.

Higher values indicate greater intra-regional inequality in accessibility.

### Multiple regression analysis

VIF values were below 5 for all variables in both models (maximum VIF = 4.08, mean VIF = 2.23), confirming the absence of substantial multicollinearity. Table 2 presents the results of the multiple regression analyses based on the Penchansky and Thomas five-dimensional access framework.

**Table 2.**
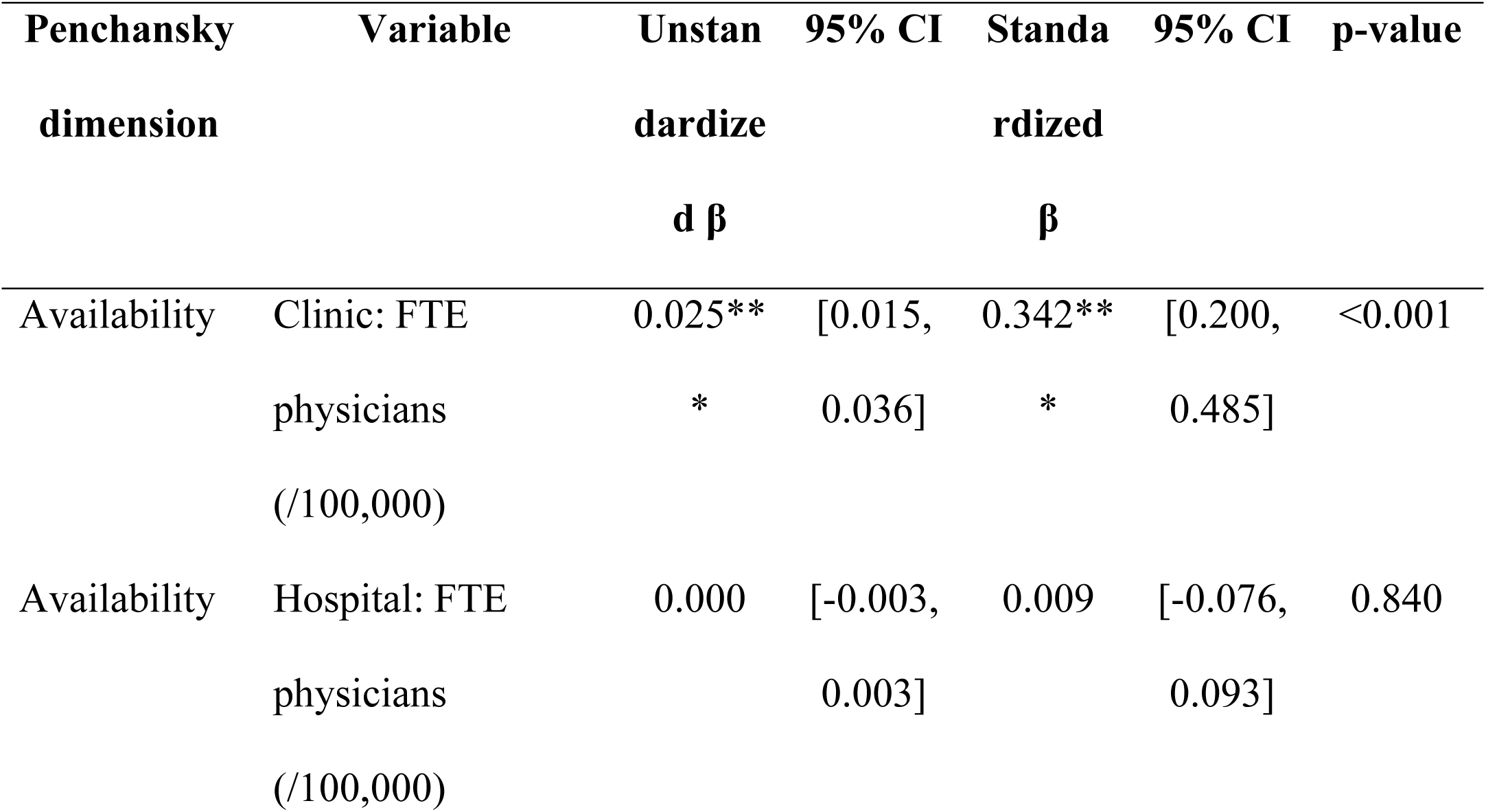
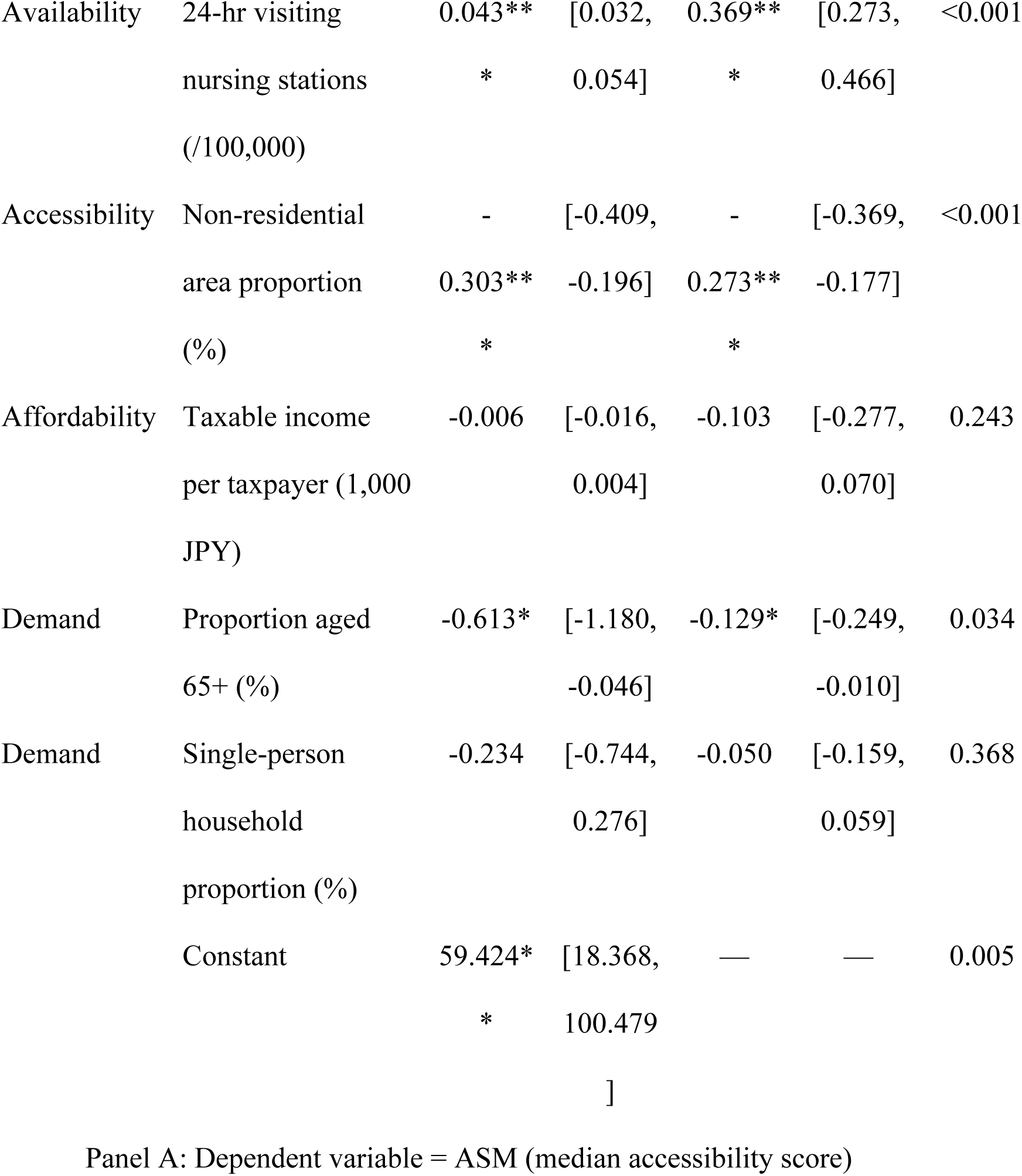
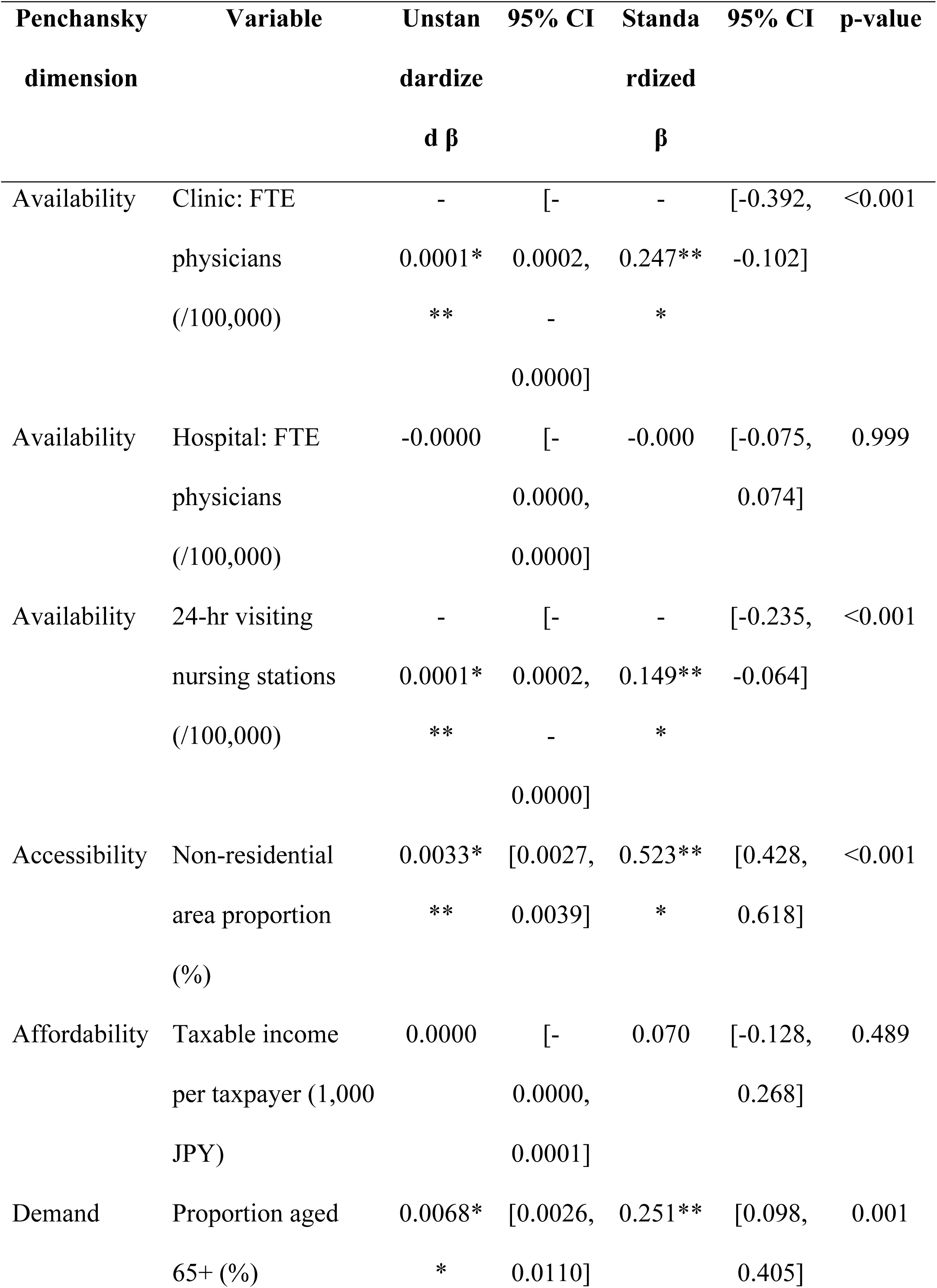
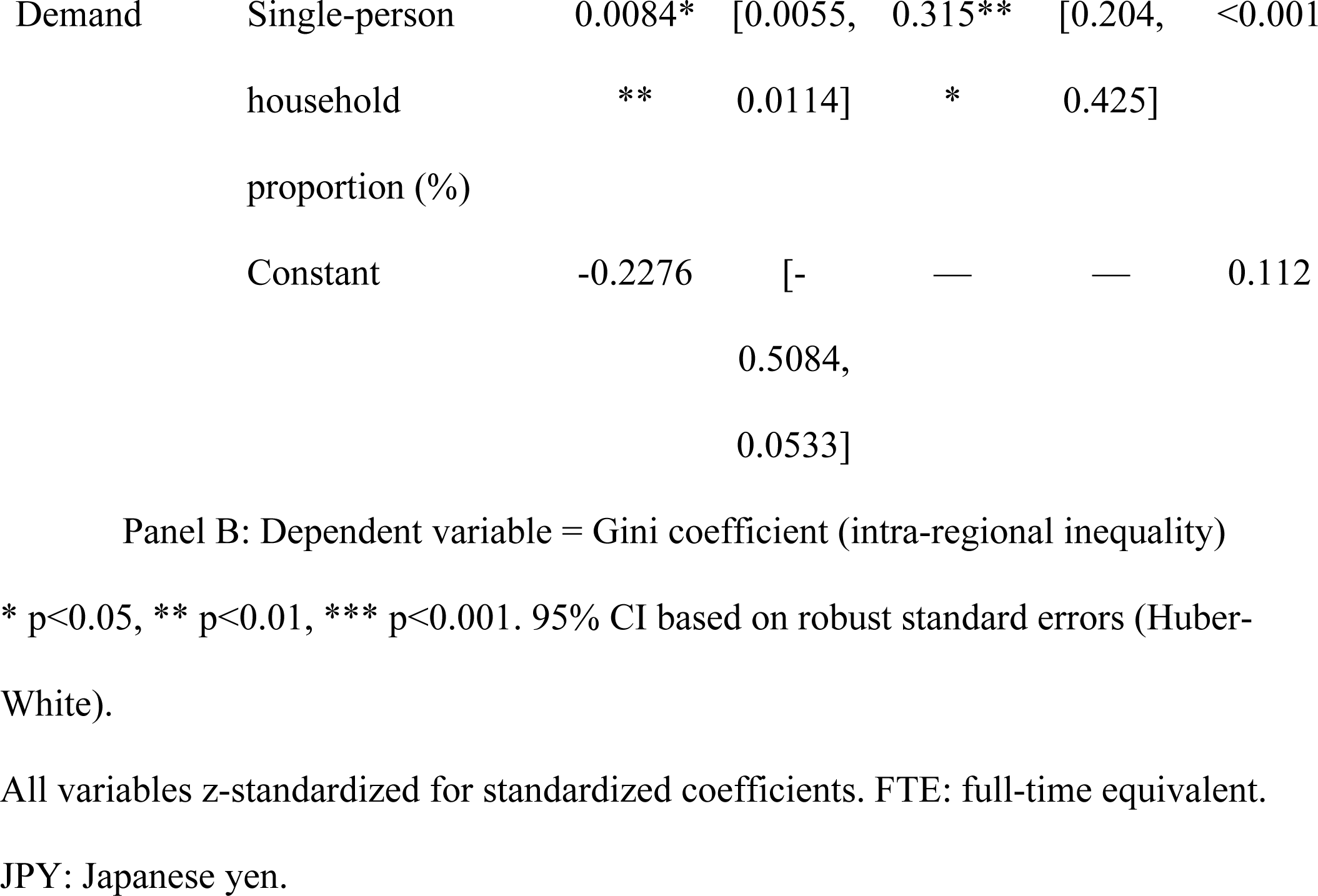
Multiple regression results (OLS, robust standard errors).

In the model with ASM as the dependent variable, variables in the Availability dimension showed the strongest associations. The number of 24-hour visiting nursing stations (β = +0.369) and the number of full-time equivalent clinic physicians (β = +0.342) were both significantly and positively associated with ASM, indicating that the availability of home healthcare supply resources was the primary determinant of accessibility. The Accessibility dimension variable, the proportion of non-residential areas (β = −0.273), showed a significant negative association, confirming the impact of geographic constraints. In the Demand dimension, the proportion of the population aged 65 years and older (β = −0.129) showed a significant negative association; however, its effect size was small relative to other variables, and the underlying mechanism cannot be determined from these data alone. The Affordability dimension variable, taxable income per taxpayer, as well as the proportion of single-person households and the number of full-time equivalent hospital physicians, were not significantly associated with ASM (Table 2, Panel A).

In the model with the Gini coefficient as the dependent variable, the structure of associated factors differed from that of the ASM model. The strongest determinant was the proportion of non-residential areas (β = +0.523) in the Accessibility dimension, indicating that geographic constraints were the primary drivers of intra-regional inequality. In the Demand dimension, the proportion of single-person households (β = +0.315) and the proportion of the population aged 65 years and older (β = +0.251) were both significantly and positively associated with greater inequality. In the Availability dimension, the number of full-time equivalent clinic physicians (β = −0.247) and the number of 24-hour visiting nursing stations (β = −0.149) were significantly and negatively associated with the Gini coefficient, indicating that greater supply resources were associated with reduced inequality. Taxable income per taxpayer and the number of full-time equivalent hospital physicians were not significantly associated with the Gini coefficient (Table 2, Panel B).

### Subgroup analysis

The CH index was maximized at k = 6, and six clusters were identified. Ranked in descending order of ASM, these were C1 (high access, equitable; n = 48, median ASM 90.7, Gini 0.25), C2 (moderate access, equitable; n = 59, ASM 62.7, Gini 0.19), C3 (moderate access, moderate inequality; n = 95, ASM 54.2, Gini 0.33), C4 (low access, moderate inequality; n = 37, ASM 28.4, Gini 0.31), C5 (low access, high inequality; n = 73, ASM 25.0, Gini 0.47), and C6 (access desert; n = 23, ASM 0.0, Gini 0.74). C6 had a median ASM of 0.00, representing areas in which access to 24-hour home healthcare within a 30-minute catchment area was virtually nonexistent (Table 3, Figure 3).

**Fig 3.**
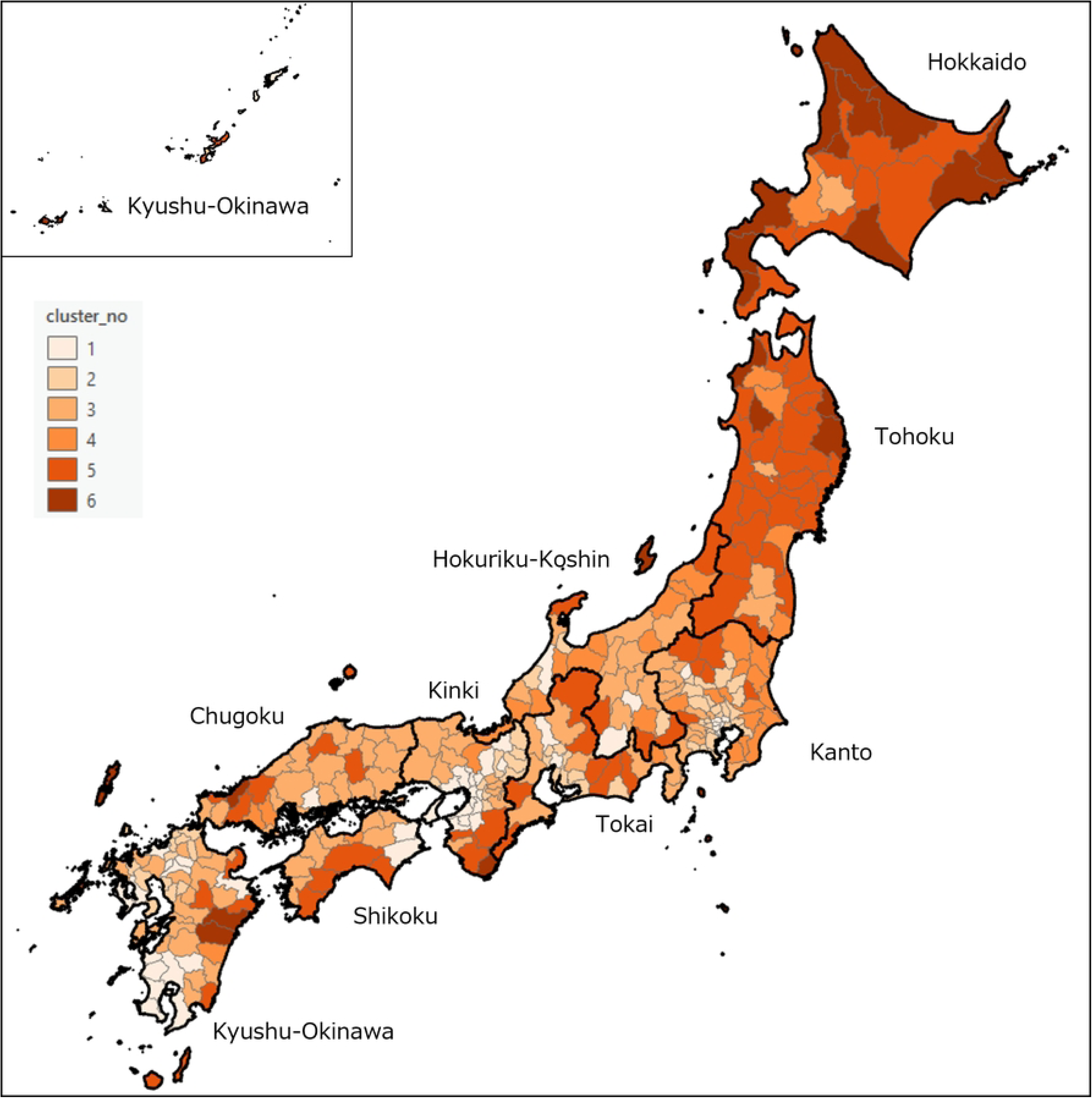
Geographic distribution of the clusters across 335 secondary medical areas.

**Table 3.**
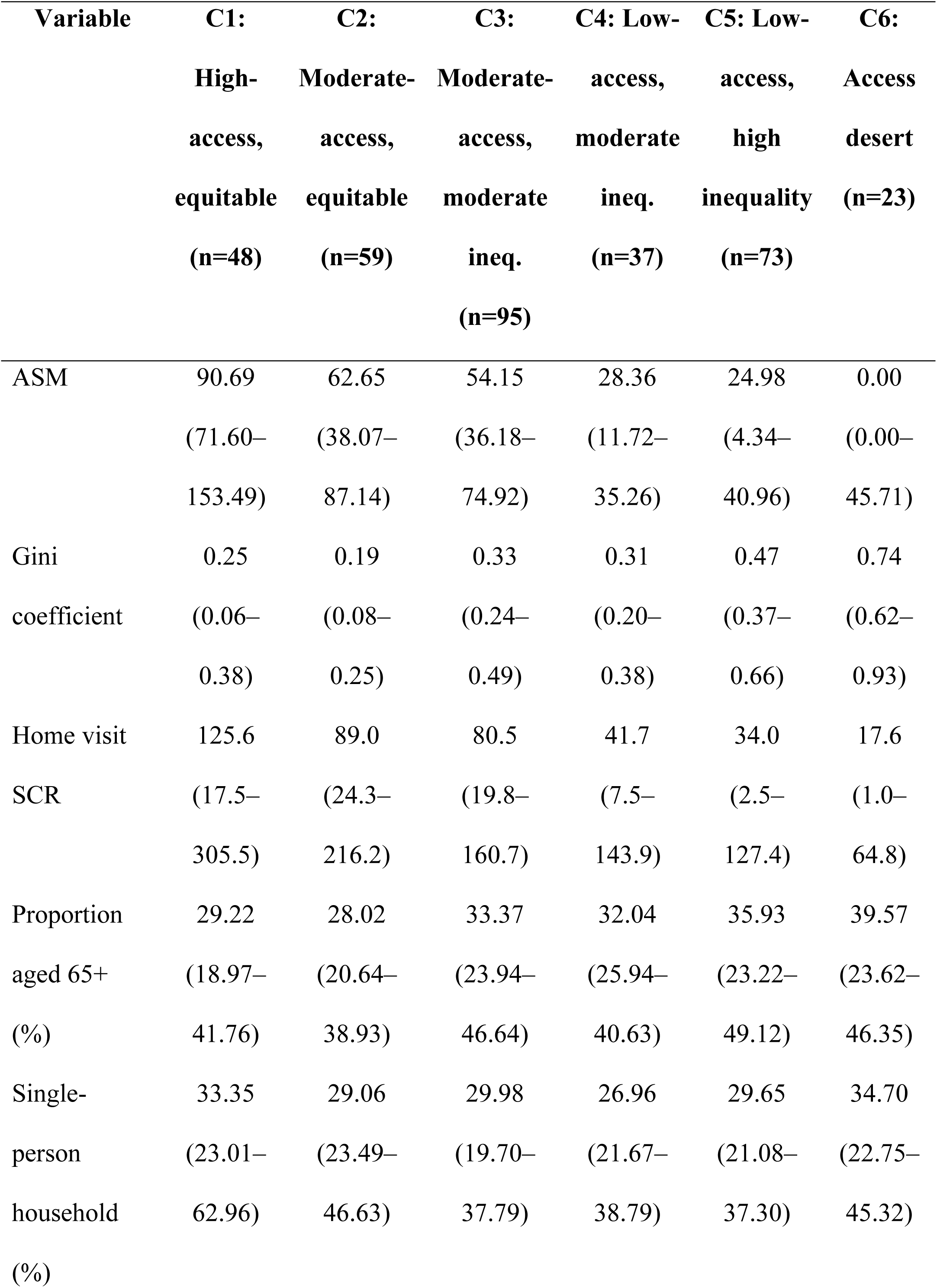
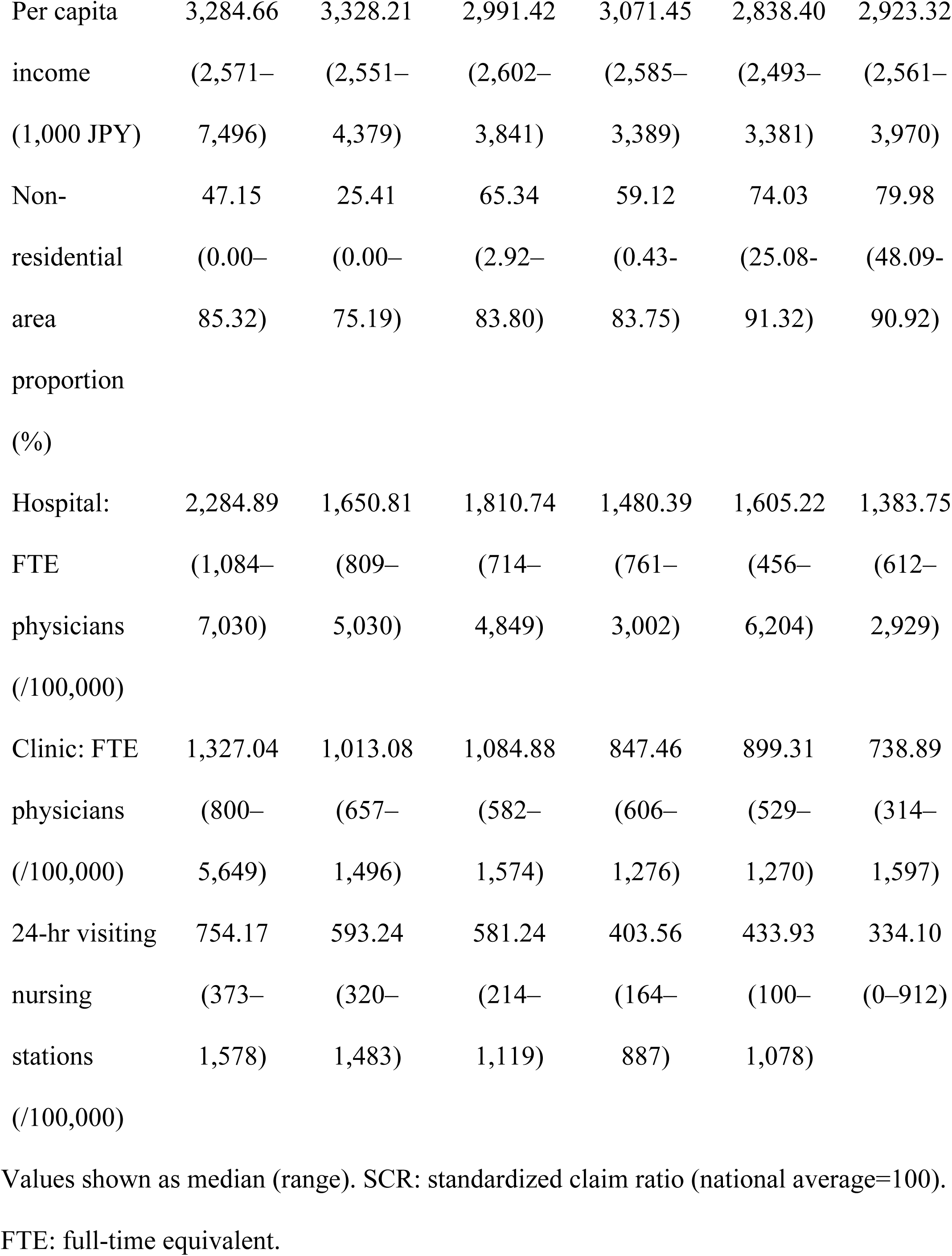
Cluster-level descriptive statistics (6 clusters, Ward’s method, k=6).

The median home-visit SCR, used as an external criterion, declined monotonically as follows: C1 = 125.6, C2 = 89.0, C3 = 80.5, C4 = 41.7, C5 = 34.0, and C6 = 17.6. The SCR, which was not included in the cluster analysis, was highly consistent with the cluster classification. The median SCR of 17.6 for C6 represents approximately 18% of the national average, indicating structurally constrained home healthcare utilization. The upper clusters (C1 and C2) had SCR values at or above the national average, whereas the lower clusters (C5 and C6) were below one-third of the national average.

The monotonic decline in SCR across clusters demonstrates that the six-cluster typology, constructed solely from ASM and Gini coefficients, was consistent with an external indicator (home-visit SCR) that was not included in the clustering procedure, thereby supporting external validity. Notably, C2 and C3 had similar ASM values (62.7 vs. 54.2) but clearly differed in Gini coefficients (0.19 vs. 0.33), and the SCR was higher in C2 than in C3 (89.0 vs. 80.5). Similarly, C4 and C5 had comparable ASM values (28.4 vs. 25.0) but substantially different Gini coefficients (0.31 vs. 0.47), with the SCR also higher in C4 than in C5 (41.7 vs. 34.0). These patterns suggest that not only the level of access but also the degree of intra-regional equality may be reflected in actual healthcare utilization.

## Discussion

This study used the E2SFCA method in a nationwide analysis to quantitatively evaluate accessibility and regional disparities in 24-hour home healthcare. The results revealed a continuous band of areas characterized by low ASM and high intra-regional inequality extending from the Hokkaido to the Tohoku region, demonstrating substantial variation in accessibility across Japan.

### Geographic variation in accessibility and nationwide polarization

ASM was higher in metropolitan areas and markedly lower in the Hokkaido and Tohoku regions. Studies of primary care accessibility in Australia have also reported lower access in rural areas [33]; however, a distinctive finding of the present study was the clear difference between eastern and western Japan, even within rural areas. A prior study on visiting nursing in Hokkaido reported intra-prefectural disparities [11], but the present study demonstrated, through a nationwide comparison, that the Hokkaido and Tohoku regions represent the most access-challenged areas in Japan.

A similar geographic distribution was observed for Gini coefficients, with high values extending from the Hokkaido to the Tohoku region and low values in metropolitan centers. Prior research has reported community-level intra-regional disparities in accessibility [6, 34], but the present study is novel in demonstrating a strong association between Gini coefficients and the proportion of non-residential areas.

The six-cluster analysis combining ASM and Gini coefficients revealed a three-tier structure. The upper tier (C1 and C2) comprised equitable types concentrated in metropolitan areas, whereas the lower tier (C5 and C6) comprised high-inequality and access-desert types distributed continuously across the Hokkaido and Tohoku regions.

A notable finding of the six-cluster analysis was the presence of cluster pairs with similar ASM levels but clearly different Gini coefficients. C2 (moderate access, equitable) and C3 (moderate access, moderate inequality) had similar ASM values; however, C2 had the lowest Gini coefficient (0.19), whereas C3 had a moderate value (0.33). Similarly, C4 and C5 had comparable ASM values but substantially different Gini coefficients. Conventional healthcare planning has evaluated supply–demand balance at the SMA level, but the six-cluster typology identified in this study enables more precise differentiation of the challenges faced by each region through the simultaneous evaluation of inter-regional disparities (ASM) and intra-regional disparities (Gini coefficients). C6 (access desert; n = 23), with a median ASM of 0.00, represents a fundamental structural challenge that precedes efforts to improve accessibility.

### Determinants of accessibility and inequality

Multiple regression analysis revealed that the structures of associated factors differed between the ASM and Gini coefficient models.

For ASM, variables in the Availability dimension were dominant. The number of 24-hour visiting nursing stations (β = +0.369) and the number of full-time equivalent clinic physicians (β = +0.342) showed the strongest positive associations, indicating that accessibility to home healthcare was primarily determined by the availability of supply resources. The proportion of non-residential areas (β = −0.273) in the Accessibility dimension also showed a significant negative association. The proportion of the population aged 65 years and older (β = −0.129) showed a significant negative association; however, its effect size was small relative to other variables, and the underlying mechanism cannot be determined from these data.

In the Gini coefficient model, the structure of associated factors differed from that of the ASM model. The strongest determinant was the proportion of non-residential areas (β = +0.523), indicating that geographic constraints were the primary drivers of intra-regional inequality. The proportion of single-person households (β = +0.315) and the proportion of the population aged 65 years and older (β = +0.251) were also significantly and positively associated with greater inequality. In the Availability dimension, the number of full-time equivalent clinic physicians (β = −0.247) and the number of 24-hour visiting nursing stations (β = −0.149) were significantly and negatively associated with the Gini coefficient, indicating that greater availability of supply resources contributes to reduced inequality within SMAs.

Across both models, a notable finding was that the proportion of non-residential areas was significantly associated with both lower ASM (β = −0.273) and higher Gini coefficients (β = +0.523). Prior research has discussed regional variation in home healthcare access in relation to population density or urban–rural classifications. However, to our knowledge, no study has demonstrated that the proportion of non-residential areas—an indicator of geographic constraints—simultaneously determines both the level of accessibility and the degree of intra-regional inequality. Unlike population density, the proportion of non-residential areas is independent of population size and directly reflects topographic constraints such as mountains, forests, and lakes. These topographic constraints may impede access to home healthcare by affecting road network development and travel times; however, this study was cross-sectional in design and does not identify causal pathways. Verification of the relationship between the proportion of non-residential areas and road infrastructure, as well as the pathways through which these factors affect accessibility, remains a task for future research.

Taxable income per taxpayer in the Affordability dimension was not significantly associated with either outcome in either model. However, the income indicator used in this study was an area-level aggregate and does not directly measure individual ability to pay. Kim et al. (2022) reported a positive association between income and the number of home medical care utilization days at the individual level among adults aged 75 years and older using claims data; however, their definition of home medical care included facility-based visiting care, which limits direct comparison with the present study. Therefore, this result should not be interpreted as evidence that Affordability has no influence on home healthcare access. Elucidating the effects of Affordability requires future studies using individual-level data.

The number of full-time equivalent hospital physicians was not significantly associated with either outcome in either model, confirming that accessibility and inequality in home healthcare are more strongly determined by clinic and visiting nursing station resources than by hospital resources.

### Policy implications

Asian countries are undergoing various healthcare reforms in response to demographic aging. According to a comparative study by Noda et al., Japan possesses one of the most comprehensive healthcare systems globally and has implemented substantial reforms [35]. Japan’s community-based comprehensive care system, legally formalized under the revised Long-Term Care Insurance Act of 2014, is an integrated medical and care delivery model comprising five core elements—medical care, nursing care, prevention, housing, and lifestyle support—delivered within a daily living area defined as approximately 30 minutes of travel time. In this context, intensive home medical services are essential components. In addition, a key challenge is responding to the projected increase in demand toward 2040. The new regional healthcare vision projects a 62% increase in home healthcare demand. Current policy envisions converting a portion of long-term care beds to home healthcare; however, the present findings suggest that uniform nationwide implementation of this policy would be difficult. In the regression analysis, the proportion of non-residential areas showed a negative association with ASM and a positive association with the Gini coefficient. This finding indicates that in areas with large proportions of non-residential land—such as mountainous and forested regions—accessibility is low and intra-regional inequality is high. Under such geographic constraints, resource investment alone may have limited effectiveness. Therefore, hospital bed restructuring predicated on the transition to home healthcare requires careful consideration of regional geographic characteristics.

Telemedicine may represent a potential strategy to address geographic constraints, and its utility in home healthcare has been reported [36]. However, further investigation is warranted because its effectiveness may be limited by disparities in information literacy.

### Access-utilization mismatch and implications of home-visit SCR

An important finding was that C6 (access desert) had a median SCR of 17.6, approximately 18% of the national average, demonstrating that home healthcare utilization is structurally constrained in areas where access within a 30-minute catchment area is virtually nonexistent. The lower clusters (C5 and C6) had SCR values below one-third of the national average, suggesting that access barriers may directly suppress actual healthcare utilization. Furthermore, C2 and C3 constituted a pair with similar ASM values but different Gini coefficients, and the SCR was also higher in C2 (89.0) than in C3 (80.5), suggesting that intra-regional equality was reflected in utilization levels. This finding indicates that even at the same ASM level, the degree of intra-regional equity may influence utilization patterns.

### Limitations

This study has several limitations. Distance-decay weights were specified based on prior research, and their appropriateness for 24-hour home healthcare has not been empirically validated. Travel times assumed automobile or pedestrian travel, and the availability of public transportation was not considered. Supply capacity was evaluated using category-based weights reflecting facility designation standards, without accounting for variation in the actual number of physicians, years of experience, or specialization within each category. The regression results demonstrate associations and do not establish causal relationships. Furthermore, spatial autocorrelation among adjacent SMAs was not formally modeled. Although we confirmed the stability of regression coefficients using prefecture-level clustered robust standard errors, spatial regression models (e.g., spatial lag or spatial error models) were not employed. Future studies should incorporate explicit spatial modeling to account for potential spatial dependence in accessibility patterns.

The E2SFCA (patient residence-based) and SCR (healthcare facility location-based) use different spatial reference points; however, because home healthcare is fundamentally delivered over short distances, the impact of this difference is considered relatively small. OSM road network data have been used in multiple international healthcare accessibility studies; however, coverage completeness in Japan has not been systematically validated. Coverage may be less complete in remote rural areas, potentially affecting accessibility estimates in these regions. Furthermore, Accommodation (organizational aspects such as appointment systems and operating hours) and Acceptability (patient and provider acceptance) within the Penchansky and Thomas framework were excluded from the analysis because of the difficulty of quantification using existing data, thereby limiting the explanatory scope of access determinants.

Demand estimation applied age- and sex-specific adjustments using NDB open data on home-visit medical care fee claim counts. Although this approach allows incorporation of demand heterogeneity related to age composition, claim counts represent revealed demand constrained by the existing supply system. In areas where supply is limited, demand may be underestimated because lower supply leads to fewer claims, which in turn leads to lower estimated demand. This circular structure represents an inherent source of bias in the demand estimation approach.

Despite these limitations, this study is the first to analyze accessibility across approximately 430,000 points in all 335 SMAs nationwide, employing age-specific demand adjustment using NDB open data and travel-time calculations based on actual road network data to quantitatively evaluate accessibility and regional disparities in 24-hour home healthcare.

## Conclusion

This study used the E2SFCA method in a large-scale nationwide analysis to quantitatively evaluate accessibility and regional disparities in 24-hour home healthcare.

The findings suggest that policies addressing accessibility and disparities are needed in the formulation of the new regional healthcare vision and the revision of regional healthcare plans.

## Data Availability

All data underlying this study are publicly available from government and open-source repositories, as detailed in the Materials and Methods section. No new primary data were generated. The processed secondary-medical-area-level dataset used for the final analyses is provided in the Supporting Information.

## Acknowledgments

Not applicable

